# From telling to receiving: the disclosure-and-services cascade after childhood sexual abuse across 15 low- and middle-income countries

**DOI:** 10.64898/2026.11.02.26367142

**Authors:** Suruchi Sood, Gayatri Malhotra, Astha Ramaiya

## Abstract

Most children who experience sexual abuse in low- and middle-income countries never receive formal help, but few studies have followed survivors through the steps from telling someone to receiving services. In this pooled cross-sectional analysis, we combined 15 nationally representative Violence Against Children and Youth Surveys (VACS) fielded between 2013 and 2024 in sub-Saharan Africa, Latin America, and Eastern Europe to describe this disclosure-and-services cascade and the factors associated with each step.

The analytic sample comprised 3,608 respondents aged 18–24 years who reported sexual abuse before age 18 (2,852 female, 756 male). We estimated survey-weighted proportions for four steps: told anyone, knew where to seek formal help, sought help (among those who knew), and received help (among those who sought it). Design-based logistic regression with country fixed effects estimated adjusted odds ratios (aORs) for individual, family, and community factors.

Overall, 47.1% (95% confidence interval [CI] 44.4–49.7) of survivors had told anyone and 34.8% (32.2–37.5) knew where to seek formal help. Of those who knew (n = 1,208), 24.8% (20.9-29.1) sought help, and of those who sought help (n = 339), 81.9% (75.9–86.7) received it. In total, 7.0% (5.7–8.5) of survivors received formal help; this proportion ranged from 0.9% to 26.3% across countries. Compared with male survivors, female survivors were more likely to tell anyone (aOR 2.30, 95% CI 1.73–3.05) and less likely to know where to seek help (aOR 0.68, 0.50–0.93); among survivors who knew where to go, female survivors were more likely to seek help (aOR 2.66, 1.39–5.09). Knowing where to seek help was more likely with older age, more schooling, and a living mother. Survivors who had ever married or cohabited were less likely to have told anyone (aOR 0.75, 0.58–0.96). No factor other than sex was associated with seeking help among those who knew where to go.

Most survivors are lost before they reach a service: two in three do not know where to go, and three in four of those who know ultimately do not seek help. Once survivors seek help, most receive it. Interventions should make services known to young people, particularly those with less schooling, and address the barriers that keep survivors, especially young men, from using services they know about.

## Introduction

Childhood sexual abuse (CSA) is a pervasive violation of children’s rights and a global public health priority. In a 2016 analysis of population-based surveys from 96 countries, Hillis et al. [1] estimated that at least one billion children aged 2 to 17 years had experienced some form of violence in the past year. Sexual violence is a substantial part of this burden. A 2025 global analysis of data from 204 countries and territories estimated that, in 2023, the age-standardized prevalence of having experienced sexual violence before age 18, among adults aged 20 years and older, was 18.9% among females and 14.8% among males [2]. Among adolescents and young people in low- and middle-income countries (LMICs) who had experienced sexual violence, 67.3% of females and 71.9% of males had first experienced it before age 18 [2]. An earlier global meta-analysis reported a self-reported CSA prevalence of about 127 per 1,000, with higher rates among girls [3]. Sub-Saharan Africa has the highest estimated prevalence of sexual violence against boys of any world region [2].

Nationally representative Violence Against Children and Youth Surveys (VACS) also show a high burden in the region: across five sub-Saharan African countries, 16.0% of young women and 8.0% of young men reported sexual violence before age 18 [4]. Country-specific estimates of the prevalence of sexual violence before age 18 in the 15 surveys pooled in this study, from sub-Saharan Africa, Latin America, and Eastern Europe, range between 5.5% and 24.8% among females and between 1.1% and 14.8% among males (S1 Table) [5–19]. CSA violates the protections guaranteed under the Convention on the Rights of the Child [20] and is associated with long-term harms to physical and mental health [21].

Prevalence alone does not show whether survivors receive help. What determines whether a survivor is protected, supported, and able to seek justice is not only whether abuse occurs but whether it is disclosed and followed by an effective response. Disclosure is often an iterative process rather than a single event [22], and many children who experience sexual abuse do not disclose it [23]. Systematic reviews consistently identify barriers to disclosure, including shame, self-blame, fear of not being believed, fear of consequences for oneself or one’s family, and the relationship to the perpetrator, operating at the individual, family, and community levels [22–24]. In one convenience sample of 42 children and adolescents, recruited mainly through psychiatric, child-welfare, and counseling services, the reported delay averaged about 17 months and ranged from the same day to 10 years [25]. Retrospective studies of adults indicate that most children who experience abuse do not disclose the abuse during childhood [26]. Disclosure is the gateway to protection, therapeutic support, and legal redress [23,24]. However, when disclosure is delayed or absent, survivors cannot use these routes; current reviews call for earlier disclosure so that support can start sooner [22,27].

To move from documenting how often abuse occurs to identifying where the response system fails survivors, we draw on the logic of the care cascade. Developed in HIV care as the spectrum of engagement in care [28], the cascade treats help-seeking as a sequence of conditional steps. Loss at any step limits the benefit of the steps that follow, so the step with the largest loss shows where intervention is most needed. Applied to CSA, the cascade comprises four steps: disclosure (telling anyone), knowing where to seek formal help, seeking help from a formal service, and receiving help from a formal service. The VACS instrument captures each step, which makes it possible to measure how many survivors are lost along the cascade. In the 2019 Kenya survey, about one third of 18- to 24-year-olds who had experienced sexual violence in childhood knew of a place to seek help (34.8% of females and 34.2% of males), but far fewer sought help (12.5% of females and 3.2% of males) or received help (10.7% of females and 3.2% of males) [10]. To our knowledge, this cascade approach, well developed in HIV care, has not been applied to CSA at the population level across multiple LMICs.

Where survivors are lost along this cascade is unlikely to be random. Current models treat disclosure as a social-ecological process, applying Bronfenbrenner’s framework [29]: individual behavior is embedded in nested interpersonal, community, and societal systems, and disclosure is shaped by interacting individual, family, contextual, and cultural factors [22]. At the individual level, age, sex, age at first victimization, the relationship to the perpetrator, and the severity or repetition of abuse are associated with whether and to whom a child discloses. At the family level, household structure and the quality of parental relationships shape access to a trusted confidant. At the community level, social norms, stigma, and the availability of services influence whether disclosure leads to support or sanction [22,23]. Sex differences are especially salient, with evidence that boys and girls face distinct disclosure dynamics [22].

International frameworks such as INSPIRE, a package of seven strategies developed by the World Health Organization and nine partner agencies, identify response and support services as a core strategy for ending violence against children, alongside efforts to shift the social norms that inhibit disclosure [30]. However, reviews of disclosure research call for more evidence on environmental, contextual, and cultural factors [22] and for more robust study designs [23]. Even children’s own understandings of violence may differ from the categories used in survey instruments [31]. The VACS offer a rare opportunity to address this gap: standardized, nationally representative, household-based surveys fielded across many LMICs, each capturing the disclosure-and-services cascade in a comparable way. Pooling these surveys makes it possible to estimate the cascade across countries and to identify the factors associated with loss at each step with greater precision and generalizability than any single national study allows.

We pooled nationally representative VACS data from 15 LMICs to examine the CSA disclosure-and-services cascade among 18- to 24-year-olds who experienced sexual abuse before age 18. We assessed three questions: (1) what proportion of survivors tell anyone, know where to seek formal help, seek help, and receive help, overall, by country, and by sex; (2) at which step the greatest loss occurs, and whether this pattern differs across countries and by sex; and (3) which individual-, family-, and community-level factors are associated with reaching each step. By locating where survivors are lost along the cascade and identifying who is most likely to be lost, the analysis aims to inform where investment, whether in awareness, service access, or the norms that govern CSA disclosure, can most effectively reduce the gap between the occurrence of abuse and an effective response.

## Materials and Methods

### Study Design

This study is a pooled cross-sectional analysis of existing, de-identified data, and reporting follows the Strengthening the Reporting of Observational Studies in Epidemiology (STROBE) guidelines for cross-sectional studies [32] (S1 Checklist).

### Data Source: The Violence Against Children and Youth Surveys

The VACS are nationally representative, household-based surveys of females and males aged 13 to 24 years, led by national governments with technical support from the US Centers for Disease Control and Prevention (CDC) and partners, under the Together for Girls partnership [5–19]. Each survey uses a multistage cluster sampling design and measures childhood experiences of sexual, physical, and emotional violence alongside help-seeking and service receipt. Data were collected between 2013 and 2024; overall response rates ranged from 45.5% to 96.2%, and each survey completed between 788 and 8,441 interviews per sex with respondents aged 13 to 24 (S1 Table). Analyses draw on each survey’s public-use file, with the accompanying questionnaires and codebooks used to build the harmonization crosswalk described below and detailed in S2 Table.

### Survey Inclusion and Exclusion

Surveys had to meet several pre-specified criteria to be included in this analysis. The dataset had to be publicly available through Together for Girls and use the standardized VACS sexual violence and disclosure modules. The sample had to be nationally representative and household-based, drawn through multistage cluster sampling, and include both sexes aged 13 to 24 interviewed by matched-sex staff. Data collection had to occur in 2013 or later, when the VACS adopted standardized sexual violence and disclosure modules, making the surveys comparable. The file had to contain the four CSA acts with their age-at-first-event items, the four cascade items or derivable equivalents, and the sampling weight, primary sampling unit (PSU), and stratum. We excluded surveys that used pre-2013 or non-standard instruments, that sampled only sub-national or humanitarian populations, or that lacked a sexual violence cascade, and we replaced superseded older surveys with each country’s newer eligible survey.

The 15 surveys that met these criteria were Colombia (2018), Côte d’Ivoire (2018), El Salvador (2017), Eswatini (2022), Honduras (2017), Kenya (2019), Lesotho (2018), Malawi (2013), Moldova (2019), Mozambique (2019), Namibia (2019), Nigeria (2014), Tanzania (2024), Zambia (2014), and Zimbabwe (2017) (S1 Table). The questionnaire version, item numbering, and routing differ across these surveys. The differences that affect the cascade measures are described below (Outcomes; Harmonization and Derivation) and item by item in S2 Table.

### Analytic Sample

The analytic sample comprises respondents aged 18 to 24 years at interview who reported CSA before age 18. This age range follows standard VACS practice: every respondent has passed through the full period of exposure before age 18, and the recall period is short [5–19]. Of 37,086 respondents aged 18 to 24 with a sampling weight and valid CSA information, 3,799 (3,023 female and 776 male) reported being sexually abused before age 18. The sample size was set based on available public-use files and no sample size calculation was conducted.

One survey, Lesotho, required a further restriction. In 13 of the other 14 surveys (all except Moldova; see Limitations), the disclosure-and-services questions were asked of every respondent who reported sexual violence. In Lesotho, these questions were asked mainly of respondents who reported physically forced or pressured sex: 155 of the 191 survivors (81%) who reported only unwanted touching or attempted sex were not asked the disclosure-and-services questions. To keep the populations comparable, the Lesotho cascade denominator was limited to survivors of forced or pressured sex, which excluded 191 survivors. The cascade denominator therefore comprised 3,608 survivors (2,852 female and 756 male). Prevalence estimates (S1 Table) use all respondents aged 18 to 24. In all surveys, respondents aged 13 to 17 were included only in a sensitivity analysis (SA1) because their period of exposure before age 18 is incomplete.

### Measures

#### Exposure: Childhood Sexual Abuse Before Age 18

CSA is defined as any of four acts before age 18, by any perpetrator: unwanted sexual touching, attempted forced sex, pressured or coerced sex, and physically forced sex. In Honduras, El Salvador, and Moldova, the version of the pressured-sex question about partners also covers sex when the respondent was too drunk to say no. Where a survey asks a separate question about sex while too drunk to refuse, that question was not counted. An act was counted as occurring before age 18 when the reported age at the first occurrence was under 18. When the age at the first occurrence was missing, the age at the most recent occurrence was used; acts with no usable age were not counted. With this definition, our weighted prevalence estimates agree with all 30 sex-specific estimates in the published country reports to within 0.1 percentage point (S1 Table).

#### Outcomes: The Disclosure-and-Services Cascade

The cascade has four steps, each coded 0 or 1 from the harmonized items: told anyone about the experience; knew of a place to go for help, such as a health facility, the police, or social services; sought help from such a service; and received help from a service. In seven surveys, the questionnaire’s list of places also includes community or religious leaders (Eswatini, Lesotho, Namibia, and Tanzania, and in the question on receiving help in Zimbabwe), nongovernmental organizations (NGOs; Lesotho, Nigeria, and Zambia), or teachers and faith-based organizations (Nigeria and Zambia); these were kept as recorded.

The survey routing shapes how the steps can be analyzed. In most surveys, the question on seeking help is asked only of survivors who knew of a place to go, and the question on receiving help only of those who sought help. We therefore analyzed the cascade as a sequence of conditional steps: telling and knowing among all survivors, seeking help among survivors who knew where to seek help, and receiving help among survivors who sought it.

Some survivors answered no to having told anyone but reported seeking or receiving help from a service. Because a survivor cannot seek help from a service without telling someone, we counted seeking or receiving formal help as telling. This changed the value for 39 survivors in the cascade denominator and raised the pooled proportion who told anyone from 45.5% to 47.1%. In Malawi, Nigeria, Zambia, and Zimbabwe, survivors who received help were not asked the disclosure (telling anyone) question, so the self-report item was also set to 1 for them. Because of this recoding, our estimates of disclosure (telling anyone) are not directly comparable with published country values. Results based on the self-report item alone are presented as a sensitivity analysis.

#### Covariates

Covariates were limited to those that could be harmonized across the surveys (S2 Table) and were grouped by ecological level [22,29]. Individual-level covariates were sex; age in years; highest level of schooling reached (none or less than primary, primary, secondary, or higher than secondary; for respondents still in school, the level they were attending); whether the respondent had ever married or lived with a partner; lifetime physical violence by a partner, peer, parent or caregiver, or adult in the community; and lifetime emotional violence by a parent or caregiver. Family-level covariates included whether the biological mother and father were alive. Community- and peer-level covariates were whether the respondent had seen someone attacked outside the home and family before age 18, and whether the respondent talks to friends about important things. The peer item was coded 1 for a lot or some (a lot or a little in Malawi, Nigeria, and Zambia) and 0 for not very much or not at all; in Eswatini and Tanzania, it was coded 1 when the respondent agreed that they can talk about problems with friends.

Measures of community cohesion or trust, food insecurity, and closeness to parents were not asked in a comparable way in all surveys, and so were not included in the primary models; closeness to parents was examined in a sensitivity analysis. According to the El Salvador codebook, the survey did not ask the peer item, so models that include it use 14 countries. In Zimbabwe, the witnessing items were asked of a subsample only; respondents who were not asked (and 4 survivors who declined) were kept in the models with a separate indicator, which was not interpreted.

#### Survey Design Variables

Each analysis incorporates the survey design through the sampling weight, PSU, and stratum, which are present in all 15 public-use files, except that the Lesotho and Zambia files have no design stratum; district (Lesotho) and province (Zambia) were used as the stratum. PSUs were nested within strata within each country and were defined separately for females and males. In the three surveys released as separate female and male files (Malawi, Nigeria, and Zambia), strata were also defined separately by sex. The exact variable names are listed in S2 Table.

### Harmonization and Derivation

Variables were mapped to a common specification using the harmonization crosswalk, following established ex-post harmonization methods for combining surveys that measured the same constructs in different ways [33]. Each step was reduced to a common binary indicator from the corresponding summary item where one exists. The Eswatini and Tanzania surveys record help-seeking and help-receiving only through service-specific items; for these, we coded each step as 1 if the respondent reported it for any service. Mozambique uses the same instrument version but keeps summary items for seeking and receiving help. When the questionnaire routing meant that a question was not asked (for example, the question on receiving help for survivors who had not sought help), the answer was treated as not applicable rather than as missing [34]. We also report the overall proportions of all survivors who sought and who received help. In these overall rates, seeking help is 0 when the respondent did not know of a service, and receiving help is 0 when help was not sought, whereas in the conditional steps these respondents are outside the denominator. We checked every derived variable against the questionnaire and codebook for its survey.

### Statistical Analysis

All estimates account for the complex survey design (weights, strata, and PSUs), with variances estimated by Taylor-series linearization. The raw VACS weights sum to each country’s population, so pooled estimates on raw weights would be dominated by the most populous countries; Nigeria would contribute the largest share. For pooled estimates, we therefore rescaled each respondent’s weight so that the weights in each country sum to that country’s sample size. Each country then contributes in proportion to its sample, and country-specific estimates are unchanged. Pooled estimates are thus a sample-weighted average of 15 surveys fielded between 2013 and 2024, not an estimate for the population of any region.

Estimates for subgroups, such as survivors or survivors who knew where to seek help, were computed as subpopulations of the full pooled design, so that all strata and PSUs contribute to the variance estimates. We report 95% CIs on the logit scale, using the design degrees of freedom of each subpopulation. Following the reporting standard of the Demographic and Health Surveys [35], estimates based on fewer than 25 unweighted cases are suppressed and estimates based on 25 to 49 cases are shown in parentheses; the same rules apply to the country-specific estimates in S3 Table. Survivor characteristics are compared by sex with the chi-square test corrected for the survey design (Rao–Scott) [36], and mean age with a t-test that accounts for the survey design.

To identify factors associated with each modeled step (telling, knowing, and seeking help among those who knew), we fitted logistic regression models that use the survey weights and design and include an indicator for each country (country fixed effects). Because of these country indicators, we compare survivors within each country. Differences between countries, such as the number of services available, the questionnaire version used, or when the survey took place, therefore do not affect the adjusted odds ratios (aORs). We included sex as a covariate rather than stratifying the models, because only 27 male survivors sought help. No interaction terms were fitted. Age was entered as years above or below 21, the middle of the 18–24 range. Centering does not change the odds ratio for age. Models used complete cases for the outcome and all covariates (telling: n = 3,328; knowing: n = 3,350; seeking help among those who knew: n = 1,120; 14 countries). Unadjusted and country-adjusted odds ratios in the same samples are given in S4 Table. We describe receipt of help among those who sought it but do not model it, because it was high and based on few male cases.

As a supplementary analysis, we fitted multilevel logistic models that allow each country its own baseline level at each step. From these models, we estimated the intraclass correlation (ICC): the share of the variation in each step, after adjustment for covariates and on the latent (logit) scale, that lies between countries rather than between individuals. In these models, the survey weights were rescaled to sum to each country’s sample size, and each country was given the same weight [37,38]. We based our conclusions on the fixed-effects models because multilevel models need many countries to estimate their uncertainty reliably. With only 14 or 15 countries, their standard errors are likely to be too small and their estimates of between-country variation unstable [39,40]. The two approaches gave nearly identical odds ratios (S4 Table).

Because the aim is to quantify the cascade and its losses rather than to test formal hypotheses, the emphasis is on estimation (proportions and aORs with 95% CIs) rather than on null-hypothesis significance testing. Where tests are reported, α is 0.05, two-sided. We do not apply a multiplicity correction, a statistical adjustment for testing many hypotheses at once, because the cascade proportions are treated as one related set of descriptive estimates rather than as independent tests. Analyses used Stata 17 with its survey (svy) estimators. The Stata do-files and the harmonization crosswalk are provided as S1 File and S2 Table.

### Sensitivity Analyses

We repeated the models under alternative specifications in seven groups, labeled SA1 to SA7 (S4 Table). SA1 included all respondents aged 13 to 24. SA2 used complete cases for the witnessing item, which excludes Zimbabwe respondents who were not asked. SA3 replaced parental survival with closeness to the mother and to the father (13 countries; closeness is not available in the Côte d’Ivoire public-use file). SA4 was limited to survivors of forced or pressured sex; the Lesotho definition applied to all surveys. SA5 defined telling by the self-report item alone. SA6 was limited to the six surveys that distinguish lower from upper secondary schooling (Colombia, Eswatini, Lesotho, Moldova, Mozambique, and Namibia); it used the four-level schooling measure (SA6a) and a five-level measure based on the International Standard Classification of Education (ISCED; SA6b). In Namibia, respondents currently attending secondary school could not be assigned an ISCED level and were excluded from SA6a and SA6b. SA7a used the covariate set without the peer item, fitted to all 15 countries, and SA7b repeated SA7a with El Salvador’s “basic” schooling category set to missing.

### Ethical Considerations

The present study analyzes only de-identified, public-use VACS data obtained under a data-use agreement with Together for Girls. For 14 of the 15 surveys, the country report describes review of the survey protocol by the CDC Institutional Review Board and in-country review bodies, and consent and assent procedures [5–13,15–19]. The Mozambique priority indicator report mentions consent and assent but does not describe the ethics review [14]. Because the study used only de-identified public-use data, it was not submitted for institutional review board review.

### Use of Generative AI

We used Claude Opus 5.5 (Anthropic) between August and October 2026 to check the Stata code, compare survey items with source documents, format the tables, and generate Figures 1 and 2 from the Stata output without changing any values. The authors made every decision in the study. The authors ran all analyses in Stata, reviewed all code and text, and checked the numbers against the Stata output. The authors take full responsibility for this article.

**Fig 1.**
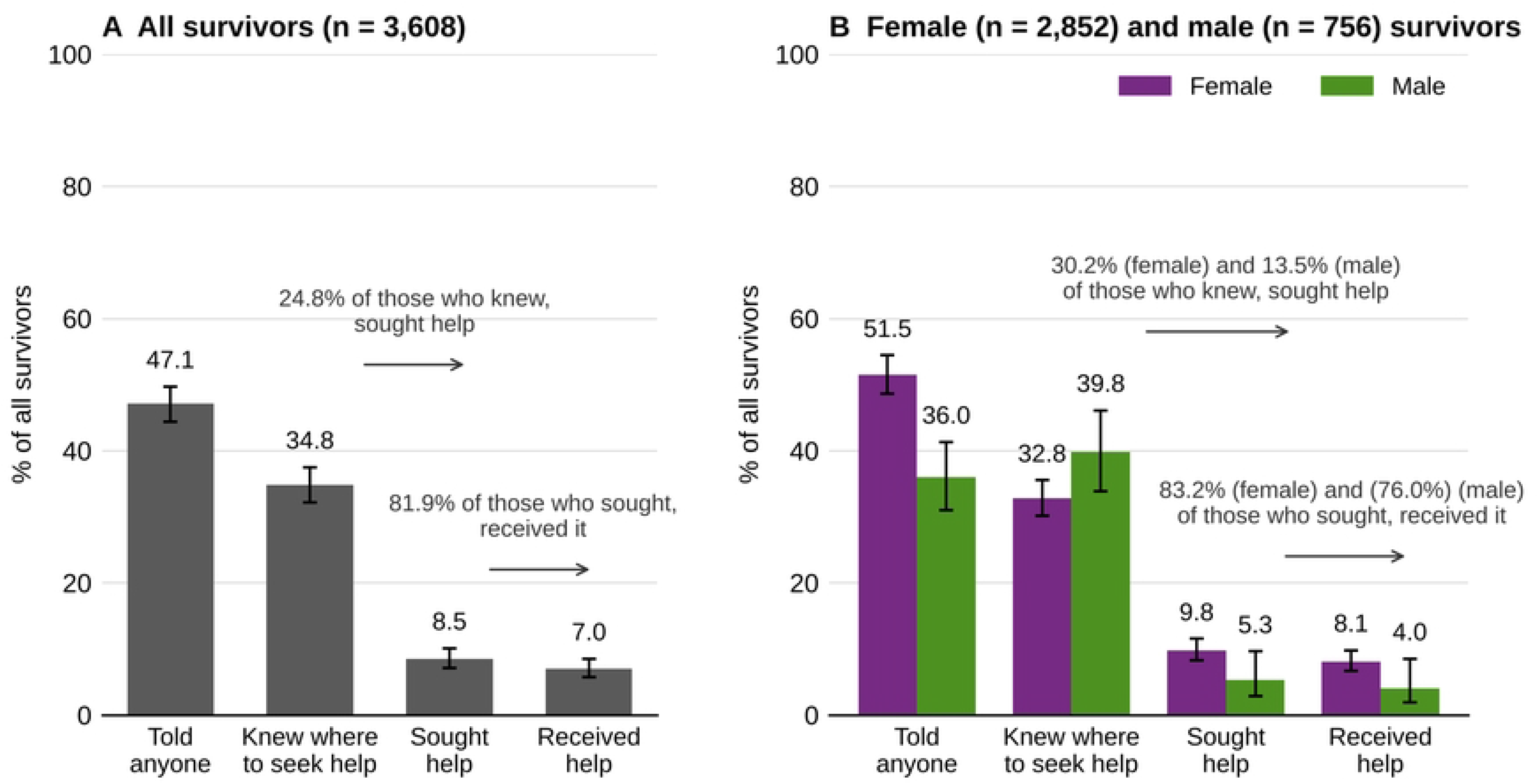
The disclosure-and-services cascade among survivors of childhood sexual abuse aged 18–24, pooled across 15 VACS surveys. (A) All survivors. (B) Female and male survivors. Bars show survey-weighted percentages of all survivors with 95% CIs, and the number above each bar gives that percentage. Arrows show the conditional steps: the percentage of survivors who knew where to seek help and sought it, and the percentage of those who sought help and received it. Values in parentheses are based on 25–49 unweighted cases. Panel n is the number of survivors in the cascade denominator; step-specific denominators exclude missing values (all survivors: 3,557 to 3,601; female: 2,812 to 2,845; male: 745 to 756). The figure was drawn with Python code written by Claude Opus 5.5 (Anthropic); all plotted values were taken unchanged from the authors’ Stata output (see Use of Generative AI).

**Fig 2.**
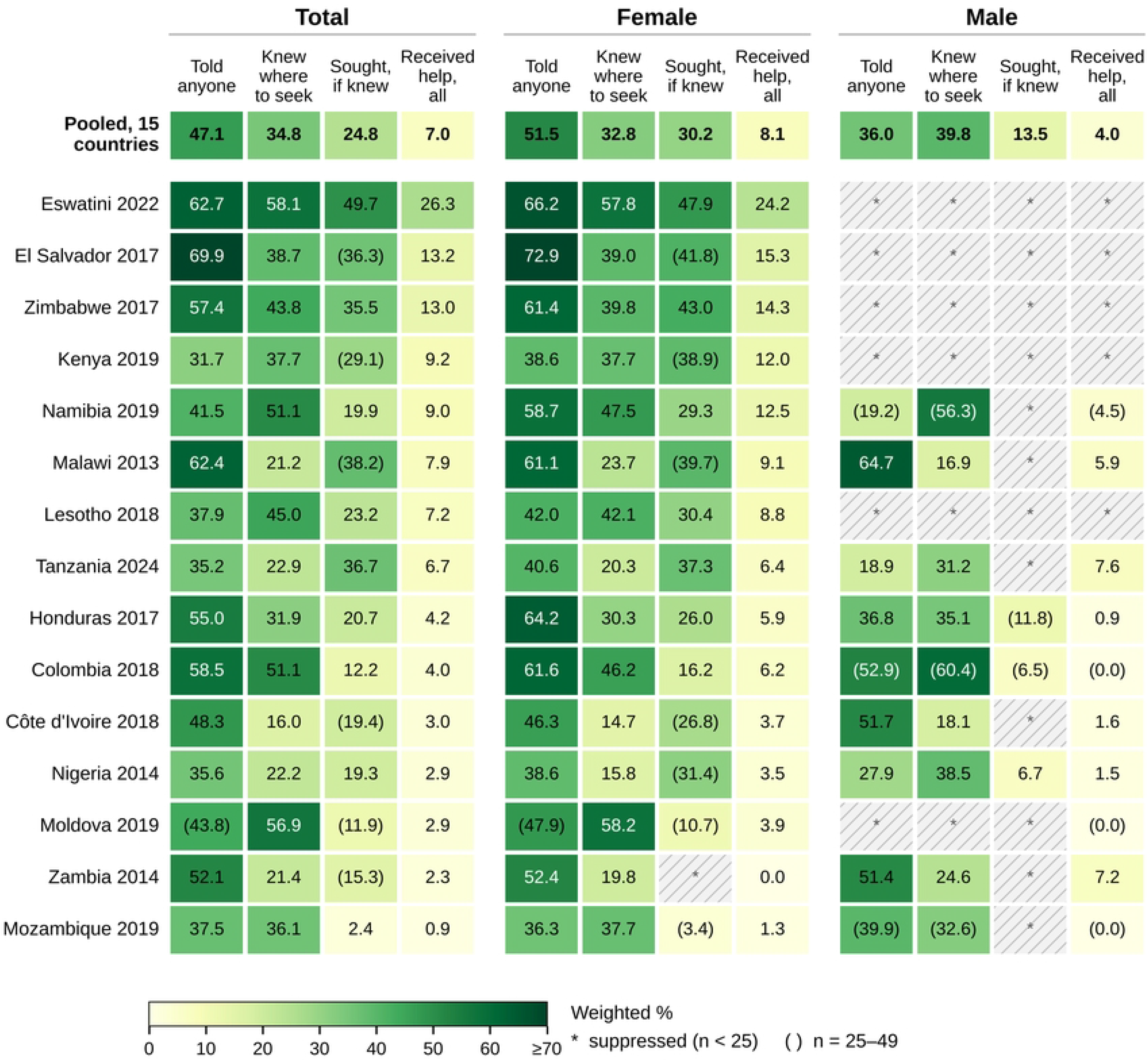
Cascade steps by survey and sex. Cells show the survey-weighted percentage of survivors who told anyone, knew where to seek help, sought help (among those who knew), and received help (among all survivors), for all survivors (Total) and for female and male survivors. Countries are ordered by the proportion of all survivors who received help, highest first. Hatched cells with * are suppressed (fewer than 25 unweighted cases); values in parentheses are based on 25–49 cases. The figure was drawn with Python code written by Claude Opus 5.5 (Anthropic); all plotted values were taken unchanged from the authors’ Stata output (see Use of Generative AI).

## Results

### Sample Characteristics

The weighted prevalence of CSA before age 18 was 13.0% (95% CI 12.3–13.7) among females and 5.7% (95% CI 5.1–6.4) among males. Across countries, it ranged from 5.5% (Eswatini) to 24.8% (Nigeria) among females and from 1.1% (Zimbabwe) to 14.8% (Malawi) among males (S1 Table).

Table 1 describes the 3,608 survivors in the cascade denominator. Their mean age was 20.4 years. Compared with male survivors, female survivors more often reported forced or pressured sex (50.5% vs 32.6%) and more often had ever married or lived with a partner (53.0% vs 20.2%). They less often reported lifetime physical violence (65.2% vs 80.6%), witnessing community violence (50.3% vs 69.6%), and talking to friends about important things (53.5% vs 73.8%; all p < 0.001). About one in three survivors of each sex reported emotional violence by a parent or caregiver (35.3% and 35.1%).

**Table 1.** Characteristics of Survivors of Childhood Sexual Abuse Aged 18–24 in the Cascade Denominator, Overall and by Sex.

| **Characteristic** | **Total, n (weighted %)** | **Female, n (weighted %)** | **Male, n (weighted %)** | **p-value, female vs male** |
|---|---|---|---|---|
| Survivors, n | 3,608 | 2,852 | 756 |  |
| **Individual** |  |  |  |  |
| Age, years, weighted mean (95% CI) | 20.4 (20.3–20.5) | 20.4 (20.3–20.6) | 20.2 (19.9–20.4) | 0.021 |
| Age 18–20 years | 2,062 (57.1) | 1,598 (55.1) | 464 (61.9) | 0.034 |
| Age 21–24 years | 1,546 (42.9) | 1,254 (44.9) | 292 (38.1) |  |
| Forced or pressured sex (vs touching or attempt only)<sup>a</sup> | 1,711 (45.4) | 1,478 (50.5) | 233 (32.6) | <0.001 |
| Schooling: none or less than primary | 257 (7.7) | 221 (8.6) | 36 (5.2) | 0.003 |
| Schooling: primary | 908 (26.3) | 747 (28.6) | 161 (20.7) |  |
| Schooling: secondary | 1,962 (52.9) | 1,526 (50.8) | 436 (58.2) |  |
| Schooling: higher than secondary | 479 (13.1) | 357 (12.0) | 122 (15.9) |  |
| Ever married or cohabited | 1,622 (43.6) | 1,458 (53.0) | 164 (20.2) | <0.001 |
| Physical violence, lifetime | 2,412 (69.6) | 1,811 (65.2) | 601 (80.6) | <0.001 |
| Emotional violence by a parent or caregiver | 1,281 (35.3) | 1,002 (35.3) | 279 (35.1) | 0.951 |
| **Family** |  |  |  |  |
| Biological mother alive | 3,079 (87.7) | 2,405 (86.7) | 674 (90.2) | 0.031 |
| Biological father alive | 2,570 (74.9) | 1,984 (73.7) | 586 (77.7) | 0.178 |
| **Community and peers** |  |  |  |  |
| Witnessed community violence: no | 1,501 (41.1) | 1,261 (45.5) | 240 (30.4) | <0.001 |
| Witnessed community violence: yes | 1,891 (55.8) | 1,382 (50.3) | 509 (69.6) |  |
| Witnessing items not asked (Zimbabwe)<sup>b</sup> | 174 (3.0) | 174 (4.2) | 0 (0.0) |  |
| Talks to friends about important things<sup>c</sup> | 2,000 (59.4) | 1,454 (53.5) | 546 (73.8) | <0.001 |

*Note.* n is unweighted; percentages are survey-weighted and exclude missing values. p values are from chi-square tests corrected for the survey design (mean age: design-adjusted t test). Missing values (n): forced or pressured sex 4; schooling 2; ever married 16; physical violence 7; emotional violence 2; mother alive 11; father alive 90; witnessing 42; talks to friends 101 (including 89 in El Salvador, where it was not asked). <sup>a</sup> Lesotho: survivors of forced or pressured sex only (see Methods). <sup>b</sup> Asked of a subsample only; this row also includes 4 survivors who declined. The p-value compares all three categories. <sup>c</sup> Not asked in El Salvador (14 countries).

### The Overall Cascade

Table 2 and Fig. 1 show the pooled cascade. Of all survivors, 47.1% (95% CI 44.4–49.7) had told anyone and 34.8% (95% CI 32.2–37.5) knew where to seek formal help. Among survivors who knew where to seek help, 24.8% (20.9–29.1) sought help, and among those who sought help, 81.9% (75.9–86.7) received it. Overall, 8.5% (7.1–10.1) of all survivors sought formal help, and 7.0% (5.7–8.5) received formal help. The largest losses occurred at the steps of knowing where to seek help and of seeking it.

**Table 2.** The Disclosure-and-Services Cascade Among Survivors of Childhood Sexual Abuse Aged 18–24, Pooled Across 15 VACS Surveys, by Sex.

| **Cascade step** | **Total, % (95% CI); events/n** | **Female, % (95% CI); events/n** | **Male, % (95% CI); events/n** |
|---|---|---|---|
| **Conditional steps** |  |  |  |
| Told anyone<br>Denominator: all survivors | 47.1 (44.4–49.7)<br>1,770/3,557 | 51.5 (48.6–54.4)<br>1,492/2,812 | 36.0 (31.0–41.3)<br>278/745 |
| Knew where to seek formal help<br>Denominator: all survivors | 34.8 (32.2–37.5)<br>1,214/3,582 | 32.8 (30.1–35.5)<br>960/2,833 | 39.8 (33.8–46.1)<br>254/749 |
| Sought formal help<br>Denominator: survivors who knew | 24.8 (20.9–29.1)<br>339/1,208 | 30.2 (25.8–35.0)<br>312/954 | 13.5 (7.4–23.5)<br>27/254 |
| Received formal help<br>Denominator: survivors who sought help | 81.9 (75.9–86.7)<br>269/339 | 83.2 (77.5–87.7)<br>250/312 | (76.0) (51.6–90.4)<br>19/27 |
| **Overall rates (denominator: all survivors)<sup>a</sup>** |  |  |  |
| Sought formal help | 8.5 (7.1–10.1)<br>339/3,601 | 9.8 (8.3–11.5)<br>312/2,845 | 5.3 (2.9–9.6)<br>27/756 |
| Received formal help | 7.0 (5.7–8.5)<br>269/3,601 | 8.1 (6.7–9.8)<br>250/2,845 | 4.0 (1.9–8.5)<br>19/756 |

*Note.* Survey-weighted percentages with 95% CIs; events/n are unweighted. Parentheses: 25–49 unweighted cases. Telling includes seeking or receiving formal help. <sup>a</sup> Survivors who did not reach the previous step count as not having sought or received help. Pooled estimates use country-normalized weights.

### Cascade by Sex

The cascade differed by sex (Table 2). Female survivors more often told anyone than male survivors (51.5% vs 36.0%) but less often knew where to seek help (32.8% vs 39.8%). Among those who knew where to seek help, 30.2% of female and 13.5% of male survivors sought it. Overall, 8.1% (6.7–9.8) of female and 4.0% (1.9–8.5) of male survivors received formal help. The estimate of receipt among male survivors who sought help (76.0%) is based on 27 cases and is therefore imprecise.

### Between-Country Variation

Cascade attainment varied widely across countries (Fig. 2; S3 Table). The proportion who told anyone ranged from 31.7% (Kenya) to 69.9% (El Salvador), and the proportion who knew where to seek help ranged from 16.0% (Côte d’Ivoire) to 58.1% (Eswatini). Among survivors who knew where to seek help, the proportion who sought help ranged from 2.4% (Mozambique) to 49.7% (Eswatini). The overall proportion who received formal help ranged from 0.9% (Mozambique) to 26.3% (Eswatini) and was below 10% in 12 of the 15 countries. In no country did more than 60% of survivors know where to seek help, or more than half of those who knew seek it. Estimates for male survivors could not be reported for five countries where fewer than 25 male survivors answered the cascade items (El Salvador, Eswatini, Kenya, Lesotho, and Zimbabwe); in Moldova, only the overall rates of seeking and receiving help could be reported for male survivors.

### Factors Associated With Each Step

Table 3 shows the adjusted odds ratios. Female survivors had higher odds than male survivors of telling anyone (aOR 2.30, 95% CI 1.73–3.05) and, among those who knew where to seek help, of seeking help (aOR 2.66, 1.39–5.09), but lower odds of knowing where to seek help (aOR 0.68, 0.50–0.93). The odds of knowing where to seek help increased with age (aOR 1.09 per year, 1.03–1.15) and with schooling (secondary vs none or less than primary: aOR 1.76, 1.04-2.96; higher than secondary: aOR 2.15, 1.17–3.97) and were higher when the survivor’s mother was alive (aOR 1.51, 1.10–2.06). Survivors who had ever married or lived with a partner had lower odds of having told anyone (aOR 0.75, 0.58–0.96), and survivors who had experienced emotional violence by a parent or caregiver had higher odds of having told anyone (aOR 1.35, 1.06–1.71). Apart from sex, no characteristic was associated with seeking help among those who knew where to go. Lifetime physical violence, the father’s survival, witnessing community violence, and talking to friends were not associated with any step.

**Table 3.**
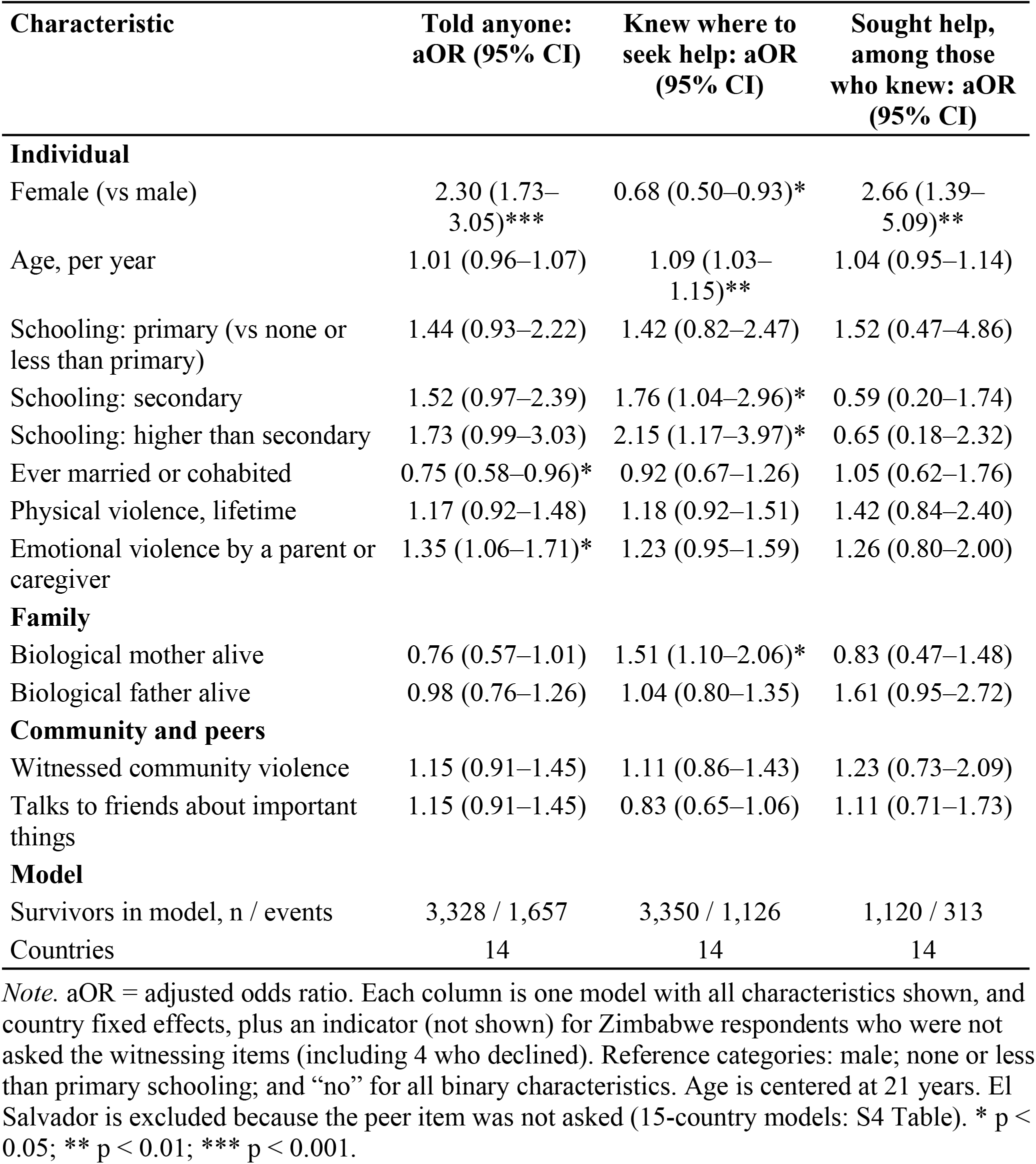
Adjusted Associations of Individual, Family, and Community Characteristics With Each Cascade Step (Design-Based Logistic Regression With Country Fixed Effects)

| Characteristic | Told anyone: aOR (95% CI) | Knew where to seek help: aOR (95% CI) | Sought help, among those who knew: aOR (95% CI) |
|---|---|---|---|
| **Individual** |  |  |  |
| Female (vs male) | 2.30 (1.73–3.05)*** | 0.68 (0.50–0.93)* | 2.66 (1.39–5.09)** |
| Age, per year | 1.01 (0.96–1.07) | 1.09 (1.03–1.15)** | 1.04 (0.95–1.14) |
| Schooling: primary (vs none or less than primary) | 1.44 (0.93–2.22) | 1.42 (0.82–2.47) | 1.52 (0.47–4.86) |
| Schooling: secondary | 1.52 (0.97–2.39) | 1.76 (1.04–2.96)* | 0.59 (0.20–1.74) |
| Schooling: higher than secondary | 1.73 (0.99–3.03) | 2.15 (1.17–3.97)* | 0.65 (0.18–2.32) |
| Ever married or cohabited | 0.75 (0.58–0.96)* | 0.92 (0.67–1.26) | 1.05 (0.62–1.76) |
| Physical violence, lifetime | 1.17 (0.92–1.48) | 1.18 (0.92–1.51) | 1.42 (0.84–2.40) |
| Emotional violence by a parent or caregiver | 1.35 (1.06–1.71)* | 1.23 (0.95–1.59) | 1.26 (0.80–2.00) |
| **Family** |  |  |  |
| Biological mother alive | 0.76 (0.57–1.01) | 1.51 (1.10–2.06)* | 0.83 (0.47–1.48) |
| Biological father alive | 0.98 (0.76–1.26) | 1.04 (0.80–1.35) | 1.61 (0.95–2.72) |
| **Community and peers** |  |  |  |
| Witnessed community violence | 1.15 (0.91–1.45) | 1.11 (0.86–1.43) | 1.23 (0.73–2.09) |
| Talks to friends about important things | 1.15 (0.91–1.45) | 0.83 (0.65–1.06) | 1.11 (0.71–1.73) |
| **Model** |  |  |  |
| Survivors in model, n / events | 3,328 / 1,657 | 3,350 / 1,126 | 1,120 / 313 |
| Countries | 14 | 14 | 14 |

*Note.* aOR = adjusted odds ratio. Each column is one model with all characteristics shown, and country fixed effects, plus an indicator (not shown) for Zimbabwe respondents who were not asked the witnessing items (including 4 who declined). Reference categories: male; none or less than primary schooling; and “no” for all binary characteristics. Age is centered at 21 years. El Salvador is excluded because the peer item was not asked (15-country models: S4 Table). \* p < 0.05; \*\* p < 0.01; \*\*\* p < 0.001.

In the multilevel models, the odds ratios were nearly identical to those in Table 3 (S4 Table). After adjustment for covariates, between-country differences accounted for 6% of the latent variation in telling, 10% in knowing where to seek help, and 18% in seeking help among those who knew (ICC 0.06, 0.10, and 0.18).

### Sensitivity Analyses

Across the nine alternative specifications (S4 Table), female survivors had higher odds than male survivors of telling anyone (aOR 2.20–2.56) and, among those who knew where to go, of seeking help (aOR 2.49–4.23); for seeking help, the 95% CI included 1 only in the six surveys that separate lower from upper secondary schooling (SA6a and SA6b). Age, schooling, and the mother’s survival were associated with knowing where to seek help in the same direction in every specification that included them, although these associations were not always statistically significant. The lower odds of knowing where to seek help among female survivors were not present among survivors of forced or pressured sex (SA4: aOR 1.05, 0.63–1.74), and the associations of telling with having ever married or cohabited and with emotional violence by a parent or caregiver were not statistically significant at ages 13 to 24, in the six-survey subsample, or among survivors of forced or pressured sex. Other associations were statistically significant in no more than three specifications (S4 Table).

## Discussion

### Principal Findings

Across 15 nationally representative surveys, fewer than half of young adults who had experienced sexual abuse in childhood had told anyone, about one in three knew where to seek formal help, and 7% had received formal help. Most survivors were lost before they reached a service: at the step of knowing where to go, and at the step from knowing to seeking help. Once survivors sought help, most received it. The pattern differed by sex. Female survivors were more likely to tell anyone and, once they knew where to seek help, to seek it. Male survivors were at least as likely to know where to go but rarely sought help. Age, schooling, and having a living mother were associated with knowing where to seek help, but no characteristic other than sex was associated with seeking it.

### Comparison With Existing Evidence

The low level of service receipt is consistent with earlier VACS analyses. Across seven countries surveyed between 2007 and 2013, Sumner et al. [41] found that 10% or fewer of survivors of childhood sexual violence received services in most countries; we found the same order of magnitude in 12 of our 15 surveys, 14 of which were conducted after 2013. The low rate is consistent with reviews of CSA disclosure, which find that many children do not disclose abuse or delay disclosure [23,27]. The cascade adds information that prevalence and disclosure rates alone do not provide. As in HIV care [28], the steps with the largest losses show where effort is most needed, although gains at one step are limited unless the later steps also improve. Here, most losses occurred before survivors reached a service rather than within the service system. The high receipt rate among survivors who sought help suggests that services usually respond when survivors reach them, although the surveys do not measure the quality of that response.

The cascade also varied widely between countries. The proportion of survivors who received formal help ranged from 0.9% in Mozambique to 26.3% in Eswatini, and after adjustment for individual characteristics, between-country differences accounted for 6% to 18% of the latent variation at each modeled step, with the largest share at the step of seeking help. The overall pattern nevertheless held in every country: in no country did more than 60% of survivors know where to seek help, or more than half of those who knew seek it. The surveys cannot explain these differences, which may reflect service availability, questionnaire version, or survey year, so comparisons between individual countries should be made with caution.

The sex differences are consistent with evidence that boys and girls face different barriers to disclosure [22]. In our data, male survivors were at least as likely as female survivors to know where to seek help, yet far fewer sought it. For males, the main barrier therefore appears to be the step from knowing to acting rather than a lack of information. This pattern is consistent with a systematic review of help-seeking for mental health problems among adolescent boys and young men, mostly in high-income countries, which identified conformity to masculine norms such as self-reliance and emotional stoicism, self-stigma, limited information about where to get help, and practical barriers such as the location, time, and cost of services [42]. The lower odds of knowing among female survivors were not present among survivors of forced or pressured sex, which suggests that this difference partly reflects differences in the type of abuse that female and male survivors reported.

The association between schooling and knowing where to seek help is consistent with schools being a source of information about services, and the association with having a living mother suggests that a parent may be a route to that information. Survivors who had ever married or lived with a partner were less likely to have told anyone. Telling may carry a higher social cost for survivors who are married or living with a partner, which could discourage disclosure, but the cross-sectional data cannot show whether the abuse, the disclosure, or the marriage came first.

### Policy and Program Implications

Three implications follow. First, services must be known: two in three survivors did not know where to seek help. INSPIRE’s response-and-support strategy calls for well-publicized, accessible mechanisms through which children can seek help [30]; such services work only if children and young people know they exist. Information delivered through schools and communities could reach many of them, and the schooling gradient suggests that young people with little schooling need other channels. Second, knowledge alone is not enough, especially for young men: three in four survivors who knew where to go did not seek help, and among male survivors nearly nine in ten did not. Services and messages may need to address confidentiality, stigma, gender norms, and the specific barriers that male survivors face. Third, because disclosure is often delayed, sometimes for many years [25–27], services and legal remedies should remain available to survivors who seek help long after the abuse, including through reforms to extend or eliminate civil and criminal statutes of limitations where necessary.

### Strengths

To our knowledge, this is the first analysis of the full CSA disclosure-and-services cascade that pools nationally representative surveys from many LMICs. The surveys share a common core instrument, and we documented item-by-item harmonization for each survey. Our prevalence estimates reproduce the published country estimates. All estimates use the full survey design. The sex difference in disclosure (telling anyone) was in the same direction in all nine sensitivity specifications, and the sex difference in seeking help in all eight that modeled it, although some other associations depended on the specification.

### Limitations

Several limitations apply. First, the data are cross-sectional and self-reported; recall and the willingness to report abuse and help-seeking may differ by sex and by country. Second, the surveys differ in year (2013 to 2024) and in instrument version, and the harmonized items are not identical across surveys. Country fixed effects remove stable differences between countries but not differences in how individual items perform. In some surveys, the list of formal services includes community or religious leaders, NGOs, or teachers; El Salvador’s “basic” schooling category (grades 1 to 9 in the questionnaire) spans primary and lower secondary levels, and in three surveys the version of the pressured-sex question about partners also covers sex while too drunk to say no. Third, in Lesotho the cascade covers only survivors of forced or pressured sex, and in Moldova the question on telling was not asked of some survivors because of a routing error noted in the country report [13]. Fourth, the cascade questions refer to any of a survivor’s experiences, not a specific incident, and the surveys do not record the order of events, such as whether telling preceded learning about services. Fifth, only 27 male survivors sought help, so estimates for this step among males are imprecise, and most male estimates could not be reported for six countries. Sixth, the pooled estimates weight countries by sample size and do not represent the population of any region. Seventh, the models used complete cases, which excluded 7% to 8% of survivors at each modeled step, including all 89 survivors in El Salvador, where the peer item was not asked; models without the peer item, fitted to all 15 surveys, gave similar results (S4 Table). Finally, the surveys do not measure when survivors first disclosed, the quality of the services received, or whether the help met survivors’ needs.

### Future Research

Data on the timing of disclosure are needed to measure delay directly. Studies of service quality, and of why survivors who know about services do not use them, especially young men, would help design better responses. Qualitative research could clarify how marriage and family structure shape disclosure. Because the VACS are repeated, future surveys could also show whether the cascade changes over time, as repeated surveys have shown for the prevalence of violence against children in Kenya [43].

## Conclusions

Across 15 countries, most young people who experienced sexual abuse in childhood never reached a formal service. The largest losses occurred before survivors reached services: most did not know where to go, and most who knew did not seek help. Telling someone is not enough: 47% of survivors told someone, but only 7% received formal help. Policies should make services known to young people, particularly those with less schooling, and reduce the barriers that keep survivors, especially young men, from using the services they know about.

## Acknowledgments

We thank the young people who took part in the Violence Against Children and Youth Surveys, and the governments of Colombia, Côte d’Ivoire, El Salvador, Eswatini, Honduras, Kenya, Lesotho, Malawi, Moldova, Mozambique, Namibia, Nigeria, Tanzania, Zambia, and Zimbabwe, which led the surveys. We also thank the US Centers for Disease Control and Prevention for technical support to the surveys and Together for Girls for making the public-use data available. The findings and conclusions in this article are those of the authors and do not necessarily represent the official position of the US Centers for Disease Control and Prevention, Together for Girls, or the governments of the participating countries.

## Declarations

### Author Contributions (CRediT)

Suruchi Sood: Conceptualization, Data curation, Formal analysis, Methodology, Writing – original draft, Writing – review & editing. Gayatri Malhotra: Conceptualization, Writing – review & editing. Astha Ramaiya: Conceptualization, Methodology, Supervision, Writing – review & editing.

### Data Availability

The VACS data are third-party data, collected by the governments of the participating countries and distributed by Together for Girls. Other researchers can request the public-use files from Together for Girls in the same way the authors did, without special access privileges. The Stata do-files that reproduce all estimates and the harmonization crosswalk are provided as S1 File and S2 Table.

The public-use VACS datasets can be requested from Together for Girls at https://www.togetherforgirls.org/en/analyzing-public-vacs-data.

## Supporting information

**S1 Table. The 15 surveys: prevalence of childhood sexual abuse and data collection.** Part A: survey-weighted percentages with 95% CIs (logit scale) and unweighted n from this study, with the published weighted national estimate from each country report. Part B: fieldwork period, overall response rates, and completed interviews by sex, mainly from the national VACS reports (interview dates for Colombia from the public-use dataset).

**S2 Table. Harmonization crosswalk.** Source items, codes, and derivation of each variable in each survey, with survey design variable names and the schooling mappings.

**S3 Table. The disclosure-and-services cascade by survey and sex.** Survey-weighted percentages with 95% CIs and unweighted denominators.

**S4 Table. Supplementary models and sensitivity analyses.** Multilevel logistic models with a country random intercept and intraclass correlations; fixed-effects and multilevel models without the peer item in all 15 surveys; unadjusted and country-adjusted odds ratios (Part A); and adjusted odds ratios under each alternative specification in sensitivity analyses SA1–SA7 (Part B).

**S1 File. Analysis code.** Three Stata do-files that reproduce all estimates from the public-use data: harmonization of the 15 surveys (01_derive_all.do), pooling (02_merge_all.do), and analysis (03_analysis.do), with instructions for running them at the top of each file.

**S1 Checklist. STROBE checklist for cross-sectional studies.**

